# Multicenter Randomized Controlled Trial of Exercise in Aortic Dissection Survivors: Rationale, Design, and Initial Hemodynamic Data

**DOI:** 10.1101/2024.03.08.24303985

**Authors:** Yasmin A. Toy, Kayla N. House, Leslie M. Boyer, Jennifer L. McNamara, Marion A. Hofmann-Bowman, Kim A. Eagle, Michelle S. Lim, Alan C. Braverman, Siddharth K. Prakash

**Affiliations:** McGovern Medical School, University of Texas Health Science Center at Houston, Houston, TX, USA; Washington University School of Medicine, St. Louis, MO, USA; University of Michigan Medical School, Ann Arbor, MI, USA

**Keywords:** exercise, thoracic aortic dissection, thoracic aortic aneurysm, heritable thoracic aortic disease, ambulatory blood pressure monitoring, hypertension

## Abstract

There are currently no evidence-based guidelines for exercise after thoracic aortic dissection (TAD), leading to highly variable recommendations that frequently lead patients to restrict their physical activities. This multicenter randomized controlled trial was intended to evaluate the safety and efficacy of a moderate intensity guided exercise program for TAD survivors. Participants were eligible for the trial if they had a Type A or Type B dissection at least 90 days before enrollment and could attend two in-person study visits. The guided exercise circuit consisted of six aerobic, isotonic, or isometric exercises that participants continued at home with virtual follow up sessions. The primary endpoint is the change in the composite anxiety and depression PROMIS-29 T-score at 12 months. Secondary endpoints include changes in grip strength, weight, 24-hr ambulatory blood pressure, and arterial biomechanical properties measured by central arterial waveform analysis. Preliminary analysis of the first 81 enrolled participants demonstrated that the guided exercise circuit was completed safely and was not associated with severe exertional hypertension, injury, or adverse cardiovascular events. At enrollment, most participants had adverse central waveform or ABPM characteristics that are associated with increased cardiovascular mortality, such as increased arterial stiffness, nocturnal hypertension, elevated pulse pressure, or blunted nocturnal dipping. Follow up of enrolled participants with longitudinal hemodynamic data to evaluate the impact of the exercise program will conclude in October 2024.

## INTRODUCTION

Thoracic aortic dissection (TAD) is a life-threatening medical emergency caused by an intimal tear in the thoracic aortic wall (1). Although advances in the surgical intervention, prophylaxis, and early recognition of TAD have improved long-term survival, many individuals with TAD face disabling, lifelong obstacles related to anxiety about sudden risk of death, re-dissection, aortic rupture, and other chronic complications (2, 3).

Observational studies have documented decreased quality of life in TAD survivors related to changes in daily activity levels and frequent anxiety or depression (3, 4). Case reports about acute aortic dissections that occurred during high intensity exercises such as weightlifting have provoked uncertainty about the safety of exercises (5–7). Weightlifting may induce extreme elevations in blood pressure due to potent pressor and Valsalva responses (8). Anxiety about dissection risk may lead TAD survivors to minimize physical activities, leading to additional deterioration of overall cardiovascular health and mental well-being (3). There is an urgent need clarify the safety and benefits of exercise (9). TAD survivors may also benefit from accessible, personalized interventions that address mental health issues (1).

Physical activities can provide synergistic benefits in combination with antihypertensive medications, reducing morbidity and mortality across a wide spectrum of cardiovascular diseases. Regular aerobic or isometric exercise is associated with a dose-dependent decrease in blood pressure, major cardiovascular outcomes, and mortality (10). On a weekly basis, at least 150 minutes of moderate aerobic exercise and 20 minutes of isometric exercise can reduce systolic blood pressure (11). Moderate aerobic activity decreased aortic medial degeneration and reduced aortic dilation in a Marfan syndrome mouse model (6). Abdominal aortic aneurysm expansion rates decreased in patients who engaged in moderate intensity exercise, especially when coupled with improved control of systolic hypertension (12). Cardiac rehabilitation proved to be safe and effective in post-surgical TAD patients, who demonstrated increased aerobic capacity and quality of life (12–14). However, only one quarter of eligible patients in the United States have access to cardiac rehabilitation programs due to social and economic barriers, depriving many patients of the proven benefits related to early guided exercise after surgery or dissection (15). Therefore, exercises that can be performed at home with inexpensive and portable equipment are needed to improve access to the benefits of cardiac rehabilitation.

Regular exercise may uniquely benefit TAD survivors by improving mental health, quality of life, and functional capacity (3). The overall goal of this study is to determine if a guided exercise program consisting of static and dynamic maneuvers that can be performed at home can decrease anxiety and increase confidence to engage in physical activities while lowering systolic blood pressure, arterial stiffness, and other cardiometabolic health measures (11). We demonstrate how this reproducible protocol can be used to improve the mental and physical well-being of TAD patients.

## METHODS

### Inclusion Criteria and Enrollment

The study protocol was reviewed and approved by the Committee for the Protection of Human Subjects at the University of Texas Health Science Center at Houston (UTHealth Houston), University of Michigan, and Washington University School of Medicine. This study was designed to proceed for 18 months: an anticipated 6 months to complete enrollment and 12 months of follow up for each participant (**Figure 1**). Aortopathy clinics at UTHealth Houston, Washington University in St. Louis, and the University of Michigan will recruit a total 126 patients (42 at each site), male and female, through clinician referral, medical records, databases, and social media campaigns. Patients who survived a thoracic aortic dissection (Type A or B) at least three months prior to study enrollment were eligible for inclusion. All potential participants were required to complete the 2009 Behavioral Risk Factor Surveillance Survey (BRFSS) about weekly time engaged in moderate and strenuous physical activities (Supplemental Text S1). Patients were excluded if any of the following apply: routine participation in greater than 150 minutes per week of moderate intensity exercises (as assessed by the BRFSS); unable to attend at least one exercise training session in person; uncontrolled hypertension (mean SBP greater than 160 mmHg at rest); symptomatic aortic, coronary, or vascular disease; unable to complete exercise program due to physical limitations, equipment or space limitations, or time commitment; do not own a treadmill or stationary cycle or have regular access to one at a gym. If patients are participating in cardiac rehabilitation, enrollment will be delayed until after discharge from the rehabilitation program.

**Figure 1.**
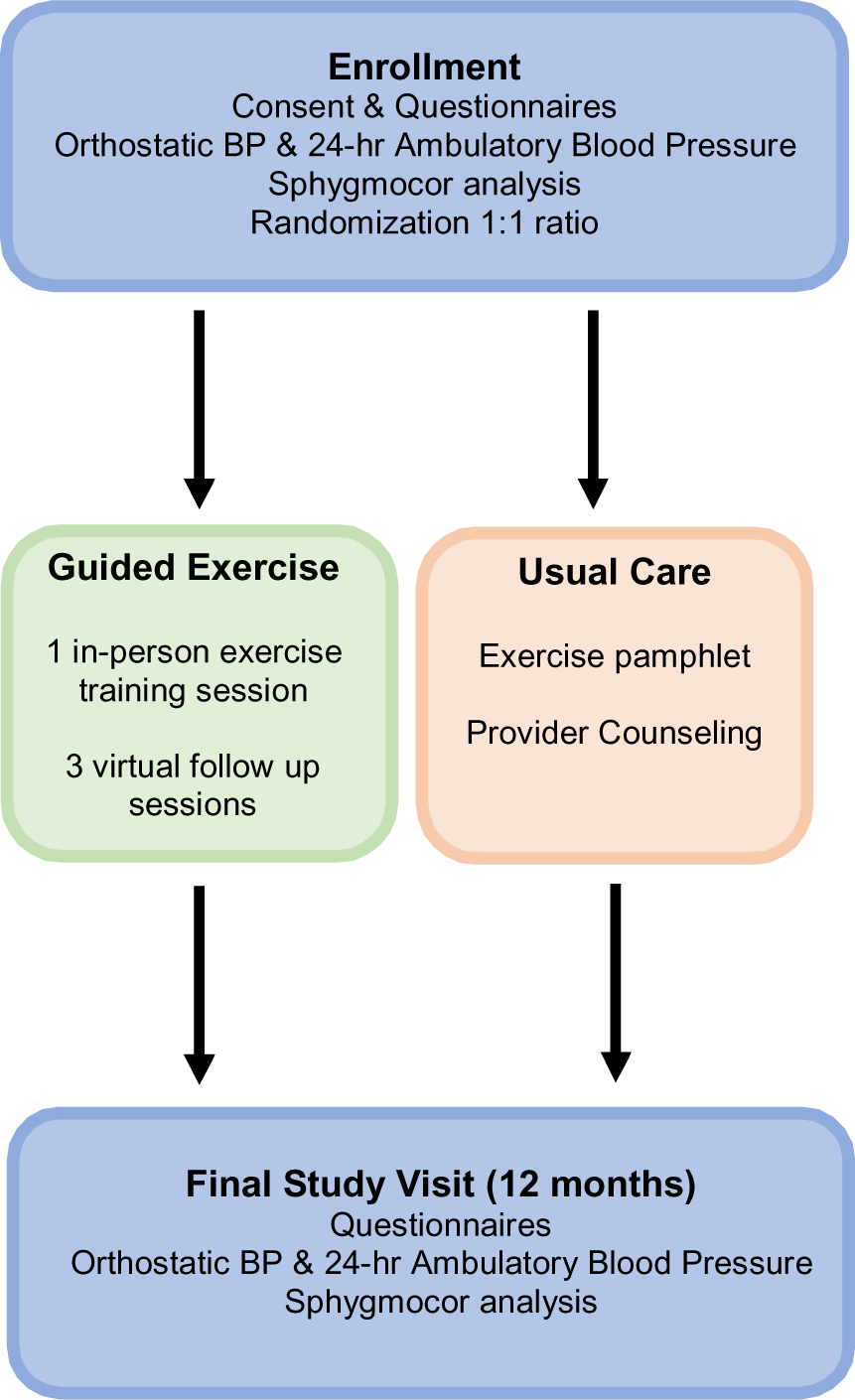
Clinical trial design and workflow for guided exercise and usual care groups. Guided exercise participants returned surveys about their routine activities and/or exercise diaries at each study visit.

After confirmation of eligibility and consent, all participants completed a demographic survey (age, sex, race, ethnicity), and the PROMIS-29 v2.0 profile questionnaire (Patient-Reported Outcomes Measurement Information System), which is validated to assess seven health domains (physical function, fatigue, pain, depressive symptoms, anxiety, ability to participate in social roles and activities, and sleep disturbance) (Supplemental Text S2). At enrollment, all participants were fitted with ambulatory blood pressure monitors to wear for 24 hours (ABPM, OnTrak, Space Labs, Inc.) with the cuff on the non-dominant arm. We performed arterial pressure waveform and pulse wave velocity analysis (Sphygmocor, AtCor Medical, Inc.) and recorded one set of orthostatic vital signs (sitting x 3, supine, standing). We also obtained consent to extract additional outcome data from health records. Participants were randomly assigned in a 1:1 ratio to receive the guided exercise program or usual care. Participants in both study arms received all usual clinically indicated care, including diagnostic tests and medications. Recommendations for tests or interventions did not change based on the assigned study arm. At the concluding study visit 12 months after enrollment, all study participants will repeat blood pressure measurements and completed the same BRFSS activity and PROMIS-29 v2.0 questionnaires as at enrollment.

### Guided Exercise

At enrollment, participants who were randomized to the guided exercise arm completed a supervised exercise protocol that included two circuits of six moderate intensity exercises: bicep curls, wall sits, hand grips, leg raises, stationary cycling and treadmill. Bicep curls were performed with the dominant hand using 5-, 8-, or 10-pound weights. Wall sits were maintained with a ninety-degree angle between the back and lower legs. Hand grip resistance level was calculated as 40% of maximal exertion using the dominant hand. Leg raises were performed in a supine position with both heels elevated six inches above ground level. Stationary cycling was performed at a target of 100 Watts. The treadmill was performed once at 3 mph at a 14% grade incline. Cardiac rehabilitation facility staff and trained study personnel supervised all exercises. Participants maintained exercises at moderate intensity (50-80% of age-adjusted maximum heart rate) long enough to acquire one brachial cuff reading (1-2 minutes). Blood pressure measurements during exercise were manually triggered using the ABPMs and supervisors ensured that the measurement arm was immobilized while the cuff inflated as recommended in the AHA scientific statement on blood pressure measurement (16). All exercises were initiated for 15 seconds prior to triggering the ABPM and maintained until the readings were completed. Post-exercise blood pressure measurements were taken following each exercise. Exertional hypertension was defined as any systolic blood pressure >180 mmHg or diastolic blood pressure > 100 mmHg on more than 1 exercise. Exercises were promptly terminated if any of the following occurred: persistent systolic pressure > 160 mmHg persisting after 3 minutes of recovery, any single systolic pressure > 210 mmHg, or any single diastolic blood pressure > 120 mmHg; chest pain, dyspnea, or significant fatigue; or a request to stop. Perceived exertion during each exercise was measured using the Borg CR-10 scale, with a score of one representing minimal exertion and a score of ten indicating maximal exertion (17). Participants received individualized instruction about how to implement the exercise program at home, with the weekly target of 5 days or at least 150 total minutes of moderate-intensity exercise. They were also counseled to record their activities in a monthly exercise diary and share fitness data recorded by home blood pressure cuffs or wearable devices.

After enrollment, the study teams followed up with participants via video check-ins and surveys. All participants completed BFRSS surveys about the intensity and frequency of their activities at 1, 3, and 9 months, and at the conclusion of the study. In the first month after enrollment, the study teams conducted one video check-in with each participant to assess any changes in health status, obtain information about clinic visits, track exercise progress, reinforce teaching about the exercise circuit, and answer questions about exercise. During the check-in, the study team also observed and corrected participants as they performed one exercise. Shorter video visits without exercise demonstrations were repeated at 3 and 9 months after enrollment, in which participants were encouraged to share their experiences with the exercise protocol and to troubleshoot potential obstacles to exercise. Participants also transmitted home blood pressure or fitness data to UTHealth if they were available. The study teams promoted the target of more than 150 minutes of moderate exercises per week at each interaction.

### Usual Care

Participants who were randomized to usual care completed 24-hour ABPM but did not receive any teaching regarding exercise and did not participate in any in-person or virtual exercise sessions.

Instead, they attended routine clinic visits and received standardized counseling about exercise, including a pamphlet with guidelines about living with aortic disease. Participants were not contacted by the study team after the initial enrollment visit.

### Data Analysis

The primary outcome is a clinically significant change in the PROMIS-29 T score or the PROMIS mental health summary score, a subset of PROMIS questions that primarily assess emotional distress (anxiety and depressive symptoms). The general population mean of PROMIS T scores is standardized at 50 points with a standard deviation of 10 points. The minimum clinically important difference (CID) is 5 points. To detect a change in 5 T score points with beta=0.80 and alpha=0.05, target sample size is 63 patients per study arm (126 total).

ABPM outcomes included mean 24-hour, daytime, and nocturnal blood pressures. Pulse pressure, nocturnal dipping status, blood pressure variability, AASI, and peak systolic pressure were also included in the analysis because they have been identified as independent predictors of cardiovascular mortality (18, 19). Study thresholds were derived from published data on ABPM norms: mean 24-hour pressure > 125/75 mmHg, mean daytime pressure > 130/80 mmHg, mean nighttime pressure > 110/65 mmHg, ambulatory arterial stiffness index (AASI) ≥ 0.70, nocturnal dipping < 10%, peak daytime systolic pressure >180 mmHg, 24-hour pulse pressure > 53 mmHg, and increased blood pressure variability, defined as a coefficient of variation > 11.1. Postural orthostasis was defined as a > 20 mm Hg decrease in systolic blood pressure and/or a > 10 mm Hg decrease in diastolic blood pressure when sitting or standing from a supine position. ABP data was analyzed using Sentinel software (v11, Space Labs, Inc., Snoqualmie, WA). Multiple comparisons were assessed using one-way ANOVA with the Tukey method.

## RESULTS

### Study cohort

A total of 445 individuals were screened and 250 were found to be eligible for the study. The major reasons why individuals were excluded were: unable to attend in-person study visits (55), exercise equipment inaccessibility (49), unable physically to exercise (42). To date, a total of 81 trial participants were enrolled with complete study data (**Table 1**). Participants who had Type A dissections (n=51) received TEVAR (n = 2), had open repairs (n = 44), or had no interventions (n = 5). Participants who had Type B dissections (n=25) received TEVAR (n = 10), had open repairs (n = 5), or had no interventions (n = 10). Participants who had more than one dissection underwent only open repairs (n = 5).

**Table 1.** Demographic Characteristics.

| Variable | All Participants<br>(n = 81) | Guided Exercise<br>(n = 38) | Usual Care<br>(n = 43) |
| --- | --- | --- | --- |
| **Age (y)** | 57 (17) | 56 (17) | 57 (19) |
| **Sex** |  |  |  |
| Female | 22 (27) | 11 (29) | 11 (26) |
| **Ethnicity** |  |  |  |
| Hispanic or Latino | 2 (2) | 1 (3) | 1 (2) |
| **Race** |  |  |  |
| American Indian/Alaska Native | 2 (3) | 0 (0) | 2 (5) |
| Asian | 5 (6) | 2 (5) | 3 (7) |
| Native Hawaiian/other Pacific Islander | 1 (1) | 0 (0) | 1 (2) |
| Black or African American | 9 (11) | 4 (11) | 5 (12) |
| White | 61 (75) | 32 (84) | 29 (67) |
| **Antihypertensive Medications** | 2 (1) | 3 (1) | 2 (1) |
| Beta blocker | 75 (93) | 37 (97) | 38 (88) |
| ACEi/ARB | 48 (59) | 25 (66) | 23 (53) |
| Diuretic | 23 (28) | 13 (34) | 10 (23) |
| Calcium channel blocker | 33 (40) | 12 (32) | 21 (48) |
| **Dissection Data** |  |  |  |
| Time since dissection (y) | 3.5 (3) | 4 (3) | 3.1 (2) |
| Type A | 51 (63) | 25 (66) | 26 (60) |
| Type B | 25 (31) | 10 (26) | 15 (35) |
| Multiple dissections | 5 (6) | 3 (8) | 2 (5) |

Values are mean (interquartile range), n (%). ACEi: angiotensin-converting enzyme inhibitor. ARB: angiotensin II receptor blockers.

### PROMIS Questionnaire

Evaluation of seven PROMIS domains found that mean T scores for anxiety (51 ± 9), pain (51 ± 7), and impairment of participation in social activities (54 ± 8) were increased. Scores for depression, fatigue, and sleep disturbance were within normal limits. There were no significant differences between PROMIS scores for participants with and without exertional hypertension, or between guided exercise and control groups.

### Grip Strength

At baseline, the mean maximum grip strength was 64 lbs (IQR 12.9). At the first follow up visit, grip strength increased by a mean of 7.8 lb (IQR 6.9).

### Orthostatic and Ambulatory Blood Pressure

At baseline, seven participants (9%) exhibited postural orthostasis. The most prevalent adverse ABPM characteristics were nocturnal hypertension (83%), blunted nocturnal dipping (40%), and elevated mean 24-hour pulse pressure (40%). Participants who developed significant exertional hypertension had higher peak blood pressure values and greater ambulatory blood pressure variability (**Table 2**). There was no association between postural orthostasis and exertional hypertension.

**Table 2.** ABPM Characteristics by Exertional Hypertension.

| Variable | Total<br>(n=71) | Exertional<br>Hypertension<br>(n=13) | No Exertional<br>Hypertension<br>(n=24) | *P* |
| --- | --- | --- | --- | --- |
| Mean SBP | 119 (16) | 122 (16) | 114 (13) | 0.13 |
| Mean DBP | 67 (12) | 70 (11) | 64 (6) | 0.06 |
| Day SBP | 123 (18) | 127 (14) | 119 (16) | 0.3 |
| Day DBP | 70 (12) | 74 (9) | 67 (5) | 0.05* |
| Night SBP | 111 (16) | 111 (19) | 103 (17) | 0.12 |
| Night DBP | 61 (14) | 62 (11) | 58 (14) | 0.22 |
| Peak daytime SBP | 157 (28) | 178 (34) | 154 (22) | 0.01* |
| Pulse Pressure | 50 (12) | 46 (10) | 48 (17) | 0.76 |
| Daytime SBP COV | 11 (4) | 14 (4) | 11 (4) | 0.03* |
| Morning Surge Index (%) | 16 (18) | 18 (28) | 18 (17) | 0.78 |
| Nocturnal Dipping (%) | 12 (12) | 17 (10) | 14 (9) | 0.10 |
| AASI | 0.53 (0.16) | 0.48 (0.08) | 0.52 (0.16) | 0.51 |

Values are mean (interquartile range). SBP: systolic blood pressure; DBP: diastolic blood pressure; PP: pulse pressure; AASI: ambulatory arterial stiffness index; Exertional hypertension: SBP > 180 mmHg or DBP > 100 mmHg in >1 exercise; COV: coefficient of variation; \*: ANOVA $P < 0.05$ .

### Safety of Exercise Protocol

All participants completed the study protocol. One in-person exercise session was temporarily delayed after a participant developed severe exertional hypertension (SBP >210), but they were able to complete the protocol after medication adjustment. Exercises that caused SBP to exceed 180 mmHg were: bicep curls (3/37, 8%), wall sits (7/35, 20%), hand grips (1/37, 3%), leg raise (1/37, 3%), stationary bicycling (4/32, 13%), and treadmill (3/36, 8%). Exercises that caused DBP to exceed 100 mmHg were: bicep curls (5/37, 14%), wall sits (11/35, 31%), hand grips (7/37, 19%), leg raise (6/37, 16%), stationary bicycling (1/32, 3%), and treadmill (3/36, 8%).

### Participant Feedback

More than half of respondents (n=23) agreed that participating in the clinical trial improved their outlook on exercise (see Supplemental Text S3 for survey details). Participants expressed increased confidence to engage in physical activities and optimism about participating in exercise after attending an in-person clinical trial visit.

### Technical Issues

Mean individual exercise completion rates from highest to lowest were bicep curls (100%), hand grip (100%), leg raise (100%), treadmill (97%), wall sit (95%), and cycle (87%). The rates for exercises with two readings per participant from highest to lowest were hand grip (92%), leg raise (92%), treadmill (87%), wall sit (84%), bicep curls (82%), and cycle (76%). The most frequent ABPM errors corresponded to excessive arm movement or vibration (Supplemental Table S1). Bracing the measurement arm successfully suppressed most of these errors [16]. We addressed the other major sources of error by changing ABPM cuff size or refitting cuffs according to manufacturer guidelines. To limit overexertion, we attempted ABPM blood pressure readings a maximum number of two times before switching to manual auscultation.

## DISCUSSION

Anxiety and uncertainty about exercise may negatively impact the cardiovascular and mental health of TAD survivors by leading them to restrict their activities. In contrast to case reports that inform current guideline recommendations, this pilot study is the first randomized controlled trial of exercise in TAD survivors. The unique objectives of this study are to assess the effects of an at-home exercise program on hemodynamic and mental health outcomes. The primary outcome is a clinically significant change in the PROMIS-29 summary T-score or mental health summary score. Secondary outcomes include the change in the burden of ambulatory hypertension and nocturnal dipping as assessed by ambulatory blood pressure monitoring. The guided exercise program proved to be safe for trial participants, and we found that grip strength, a significant predictor of cardiovascular death, increased by 30% in the first three months of participation (20, 21). We also observed adverse ABPM characteristics in many participants that are associated with increased cardiovascular mortality, such as nocturnal hypertension, blunted nocturnal dipping, or elevated pulse pressure. Ambulatory peak blood pressure and blood pressure variability predicted significant exertional hypertension. These observations highlight the high cardiovascular risk of the trial cohort. While self-reported anxiety was increased in trial participants, there was no correlation between initial PROMIS anxiety T-scores and ambulatory or exertional hypertension.

As the study progressed, we made several adjustments to home exercise instructions and the virtual visit protocol to account for the frailty and decreased physical strength of many TAD participants. The exercise instructions were altered so that participants were able to maintain moderate intensity effort without physical strain. Participants were instructed to scale up individual exercises incrementally, by increasing repetitions in 15 second increments or by two repetitions per week. When starting home exercises, we allowed participants to decrease the initial speed and incline settings of the treadmill, the angle of the wall sit, and the target rate on the stationary bicycle. The virtual visit protocol was amended to collect additional information about contacts with healthcare providers. We also provided personalized counseling to individuals who developed exertional hypertension during the in-person exercise training sessions to modify the intensity of specific exercises and to minimize Valsalva maneuvers during isometric exercises. New participants in the guided exercise study arm received the updated exercise instructions at the initial enrollment visit. Previously enrolled participants received updated instructions and teaching at virtual follow up visits.

The principal limitations to study recruitment were the requirements for participants to have access to exercise equipment at home and for in-person study visits. Technological barriers did limit timely virtual follow up visits with some participants. We plan to address these obstacles in a larger and longer trial that will be adequately powered to determine if guided exercise can reduce aortic events and prevent deaths due to TAD. In such a trial, we will collect longitudinal data on aortic enlargement, arterial stiffness, cardiac function, and serial changes in blood pressure responses to exercise over time. To promote accessibility, we plan to mail portable exercise equipment directly to participants. In the short term, we plan to adapt this protocol to create personalized exercise prescriptions for patients, and in the long-term we hope that these studies may eventually be used to develop evidence-based exercise guidelines.

## Data Availability

All data produced in the present work are contained in the manuscript or supplementary information.

## DECLARATIONS

## Acknowledgements

We are profoundly grateful to Bansari Rajani, Gabrielle Sutton, Jatin Khanna, Tara Johnson, Sue Streeter, Cardiac and Pulmonary Rehabilitation Center staff at Memorial Hermann – Texas Medical Center Hospital, and to all study participants for their time and effort. Dr. Braverman’s research is sponsored by the Pam and Ron Rubin Fund at Washington University School of Medicine and the Neidorff Aortopathy and Master Clinician in Cardiology Fellowship Program at Washington University School of Medicine. We would like to thank the staff of the cardiac rehabilitation program at Washington University School of Medicine, Barnes-Jewish Heart & Vascular Center, and the staff of the cardiac rehabilitation program at the Frankel Cardiovascular Center at University of Michigan.

## Authors’ contributions

Toy Y: Data curation, Writing - Original draft preparation; House K: Supervision, Project administration, Writing - Reviewing and Editing; Boyer L: Supervision, Project administration; McNamara J: Supervision, Project administration; Hofman-Bowman M: Conceptualization, Methodology, Writing - Reviewing and Editing, Project administration; Eagle K: Conceptualization, Methodology, Writing - Reviewing and Editing, Project administration; Braverman A: Conceptualization, Methodology, Writing - Reviewing and Editing, Project administration; Prakash S: Conceptualization, Methodology, Writing - Reviewing and Editing, Project administration.

## Availability of data and materials

Data will be published as supplementary information.

## Financial support and sponsorship

This work was supported in part by a grant from the John Ritter Foundation for Aortic Health and a gift from Carmen David.

## Conflicts of interest

All authors declared that there are no conflicts of interest.

## Ethical approval and consent to participate

The study protocol was reviewed and approved by the Committee for the Protection of Human Subjects at the University of Texas Health Science Center at Houston (HSC-MS-22-0936). All subjects signed a written informed consent document prior to enrollment.

## Consent for publication

Not applicable.

## Supplementary Material

**Supplementary Table 1.** Most frequently observed Space Labs ABPM error codes.

| Error Code | Condition |
| --- | --- |
| EC04 | Occasional EC04 messages reflect excessive patient movement.<br>Frequent EC04 messages indicate an improperly applied cuff or a monitor malfunction |
| EC10, 70, 90 | Excessive movement |
| EC11 | Monitor did not pump above mean arterial level |
| EC40 | Movement during systole |
| EC50, 58 | Movement during diastole |
| EC52 | Kinked tubing |
| EC62 | Cuff applied too loosely |

### Supplemental Text 1. BRFSS Questionnaire

#### BRFSS Questionnaire: Physical Activity

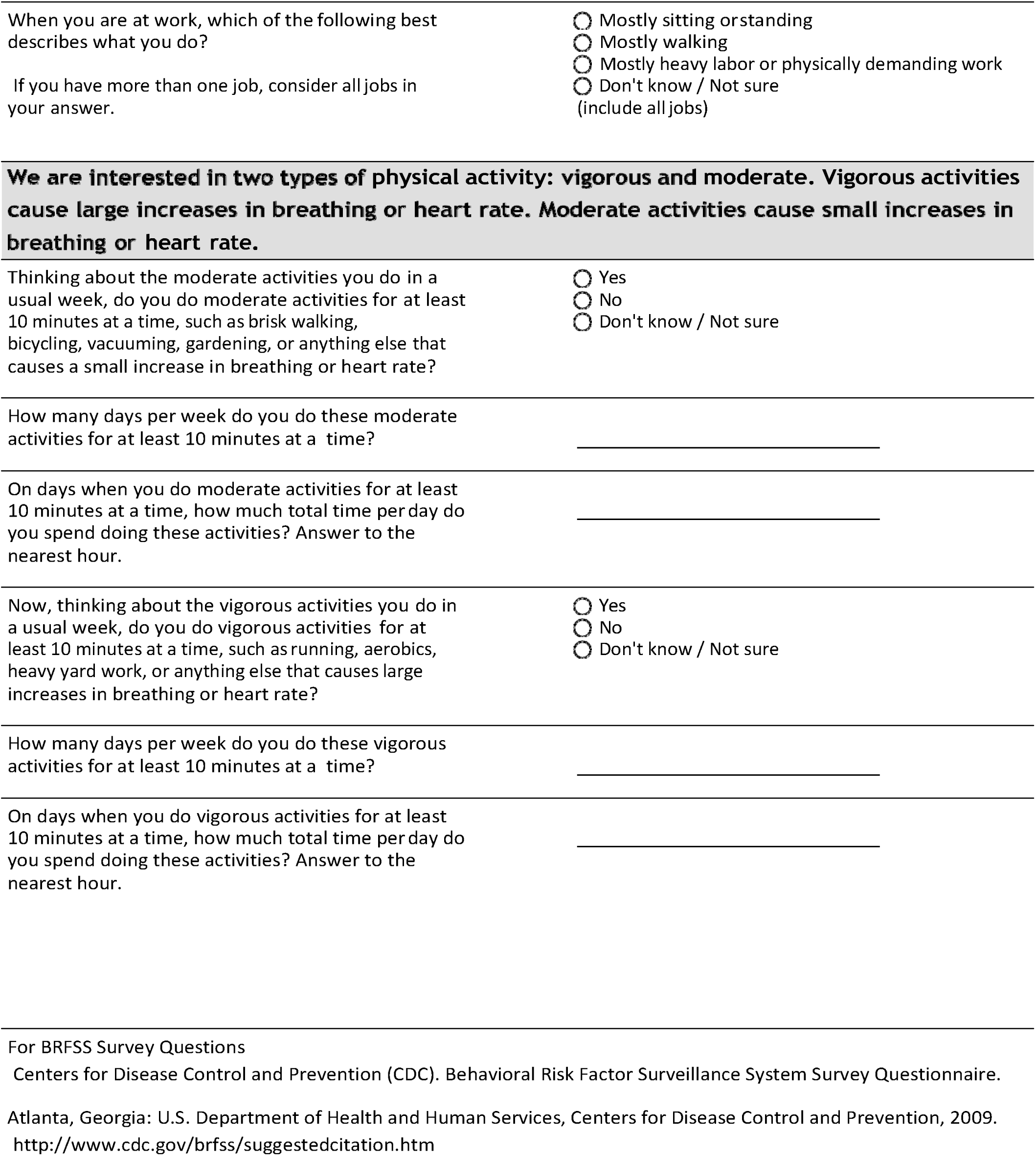

### Supplemental Text 2. PROMIS Questionnaires

#### 1) PROMIS - Physical Function

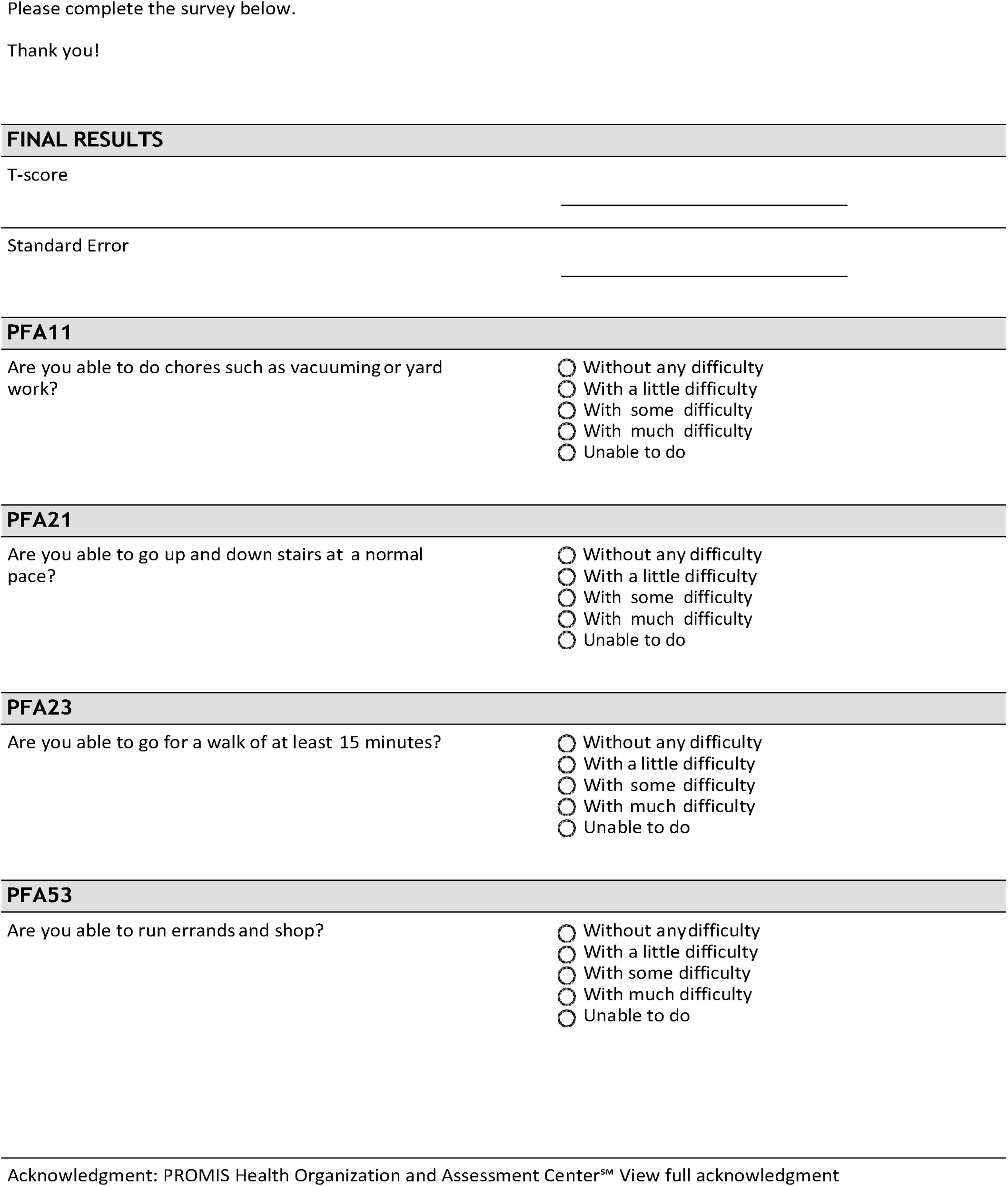

#### 2) PROMIS – Anxiety

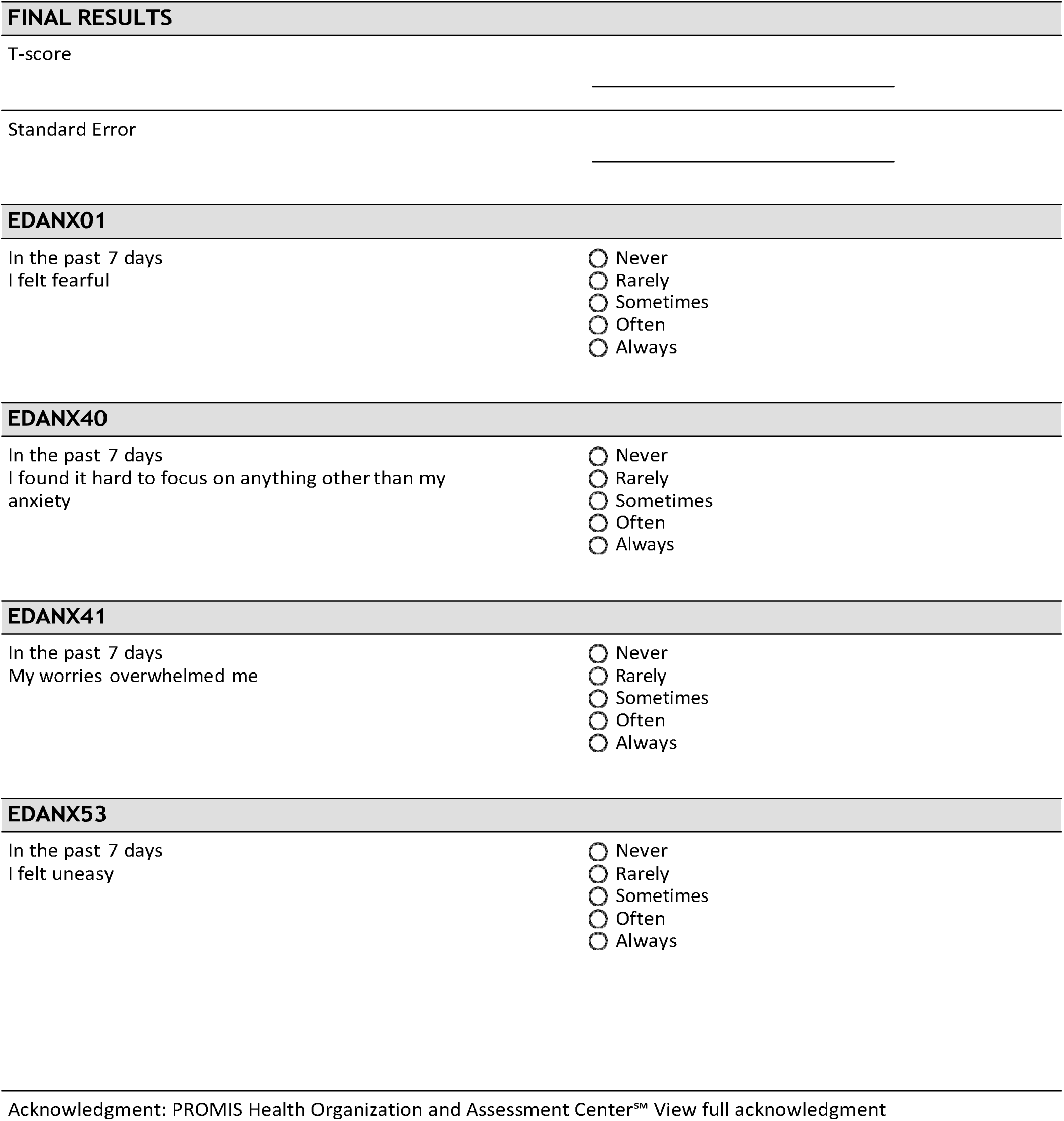

#### 3) PROMIS – Depression

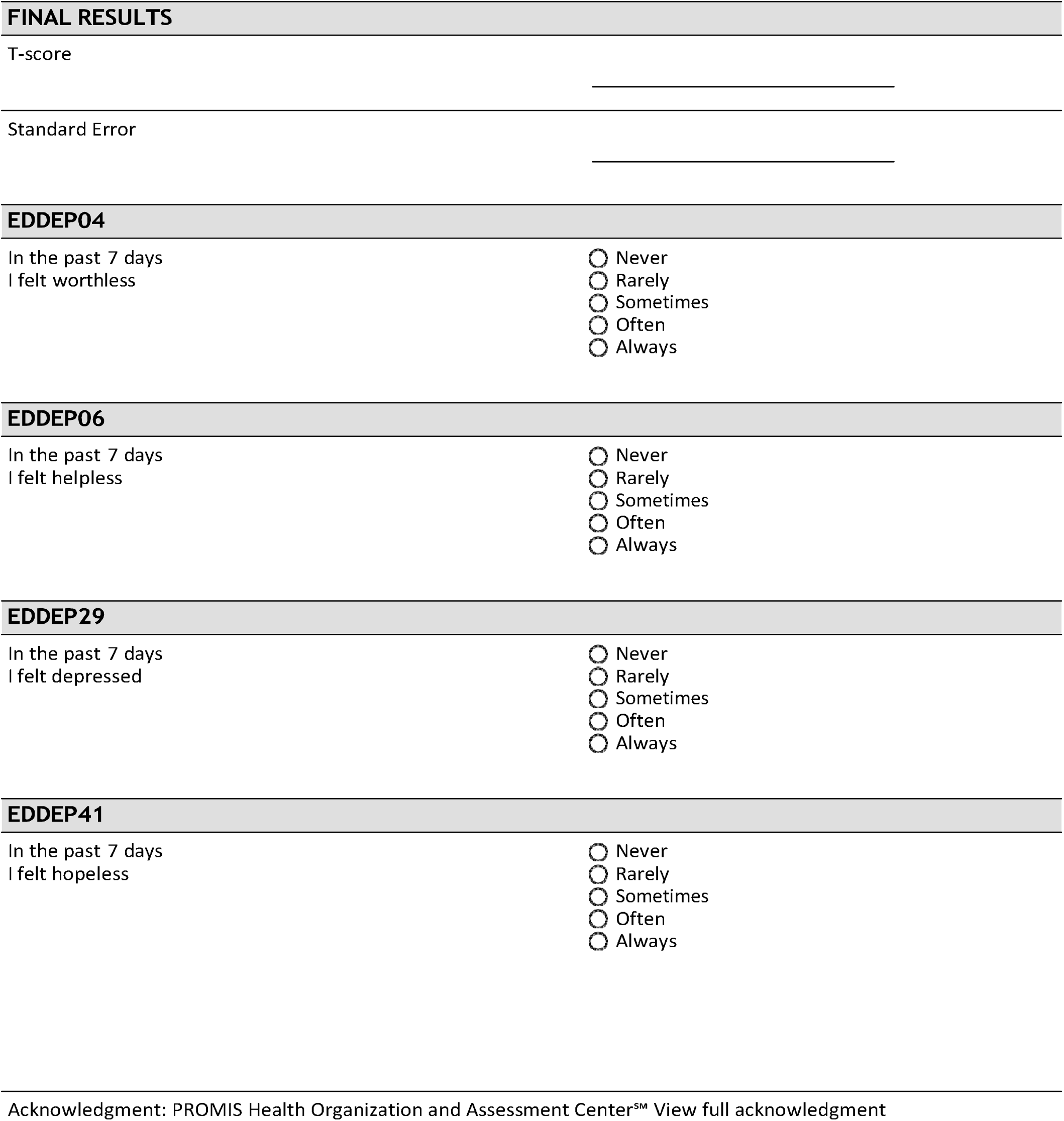

#### 4) PROMIS – Fatigue

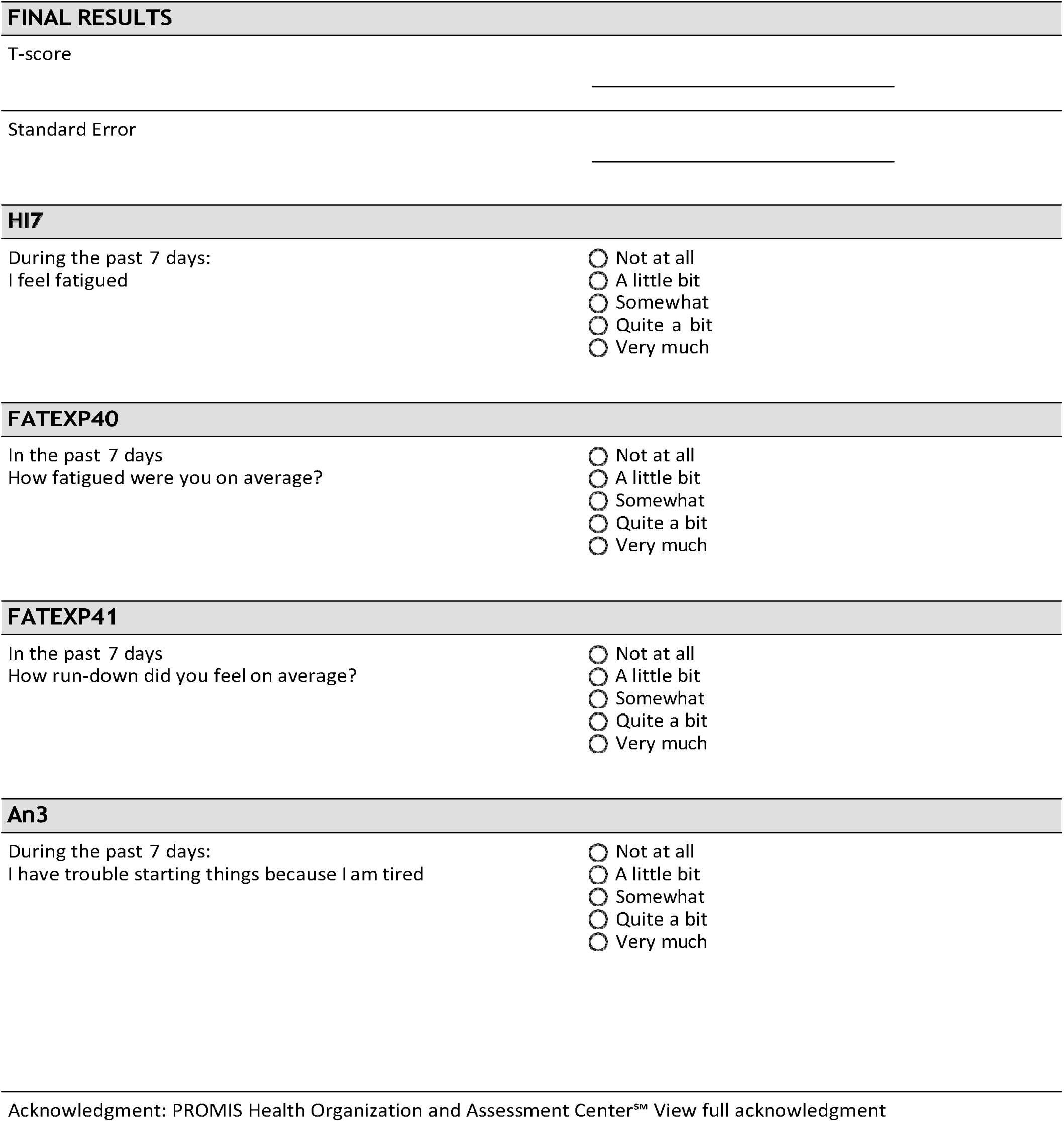

#### 5) PROMIS - Sleep Disturbance

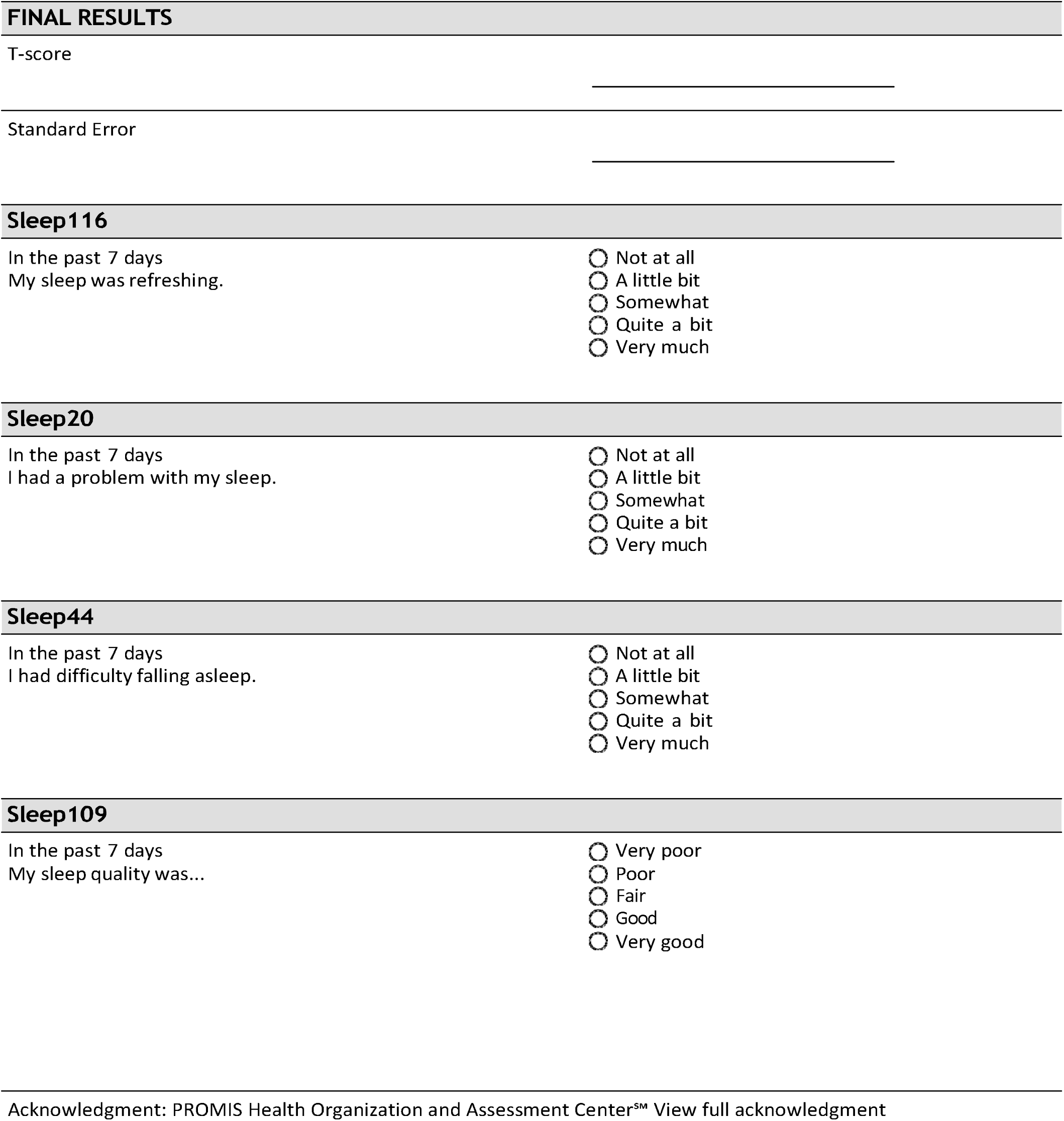

#### 6) PROMIS - Ability to Participate Social

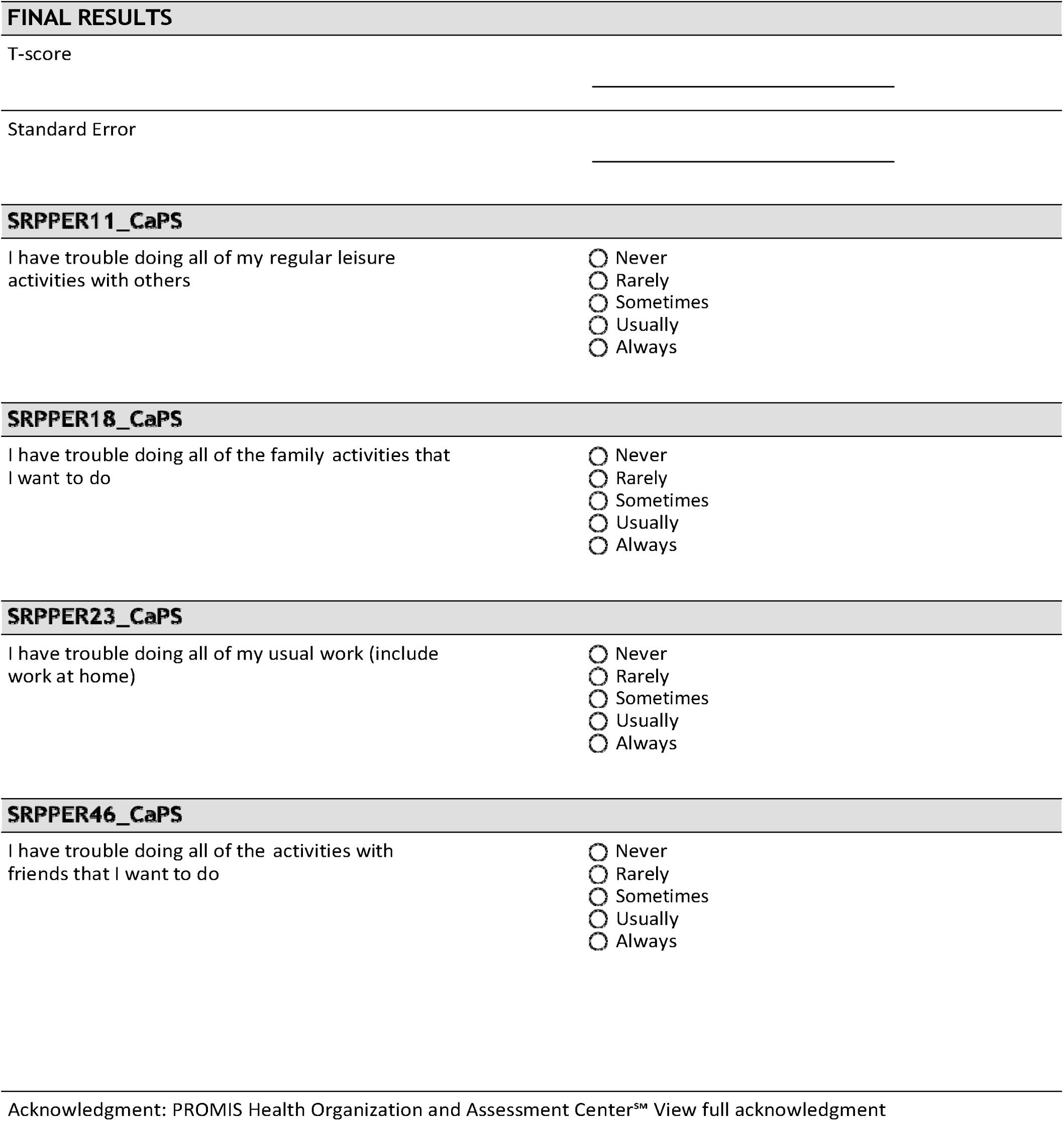

#### 7) PROMIS - Pain Interference

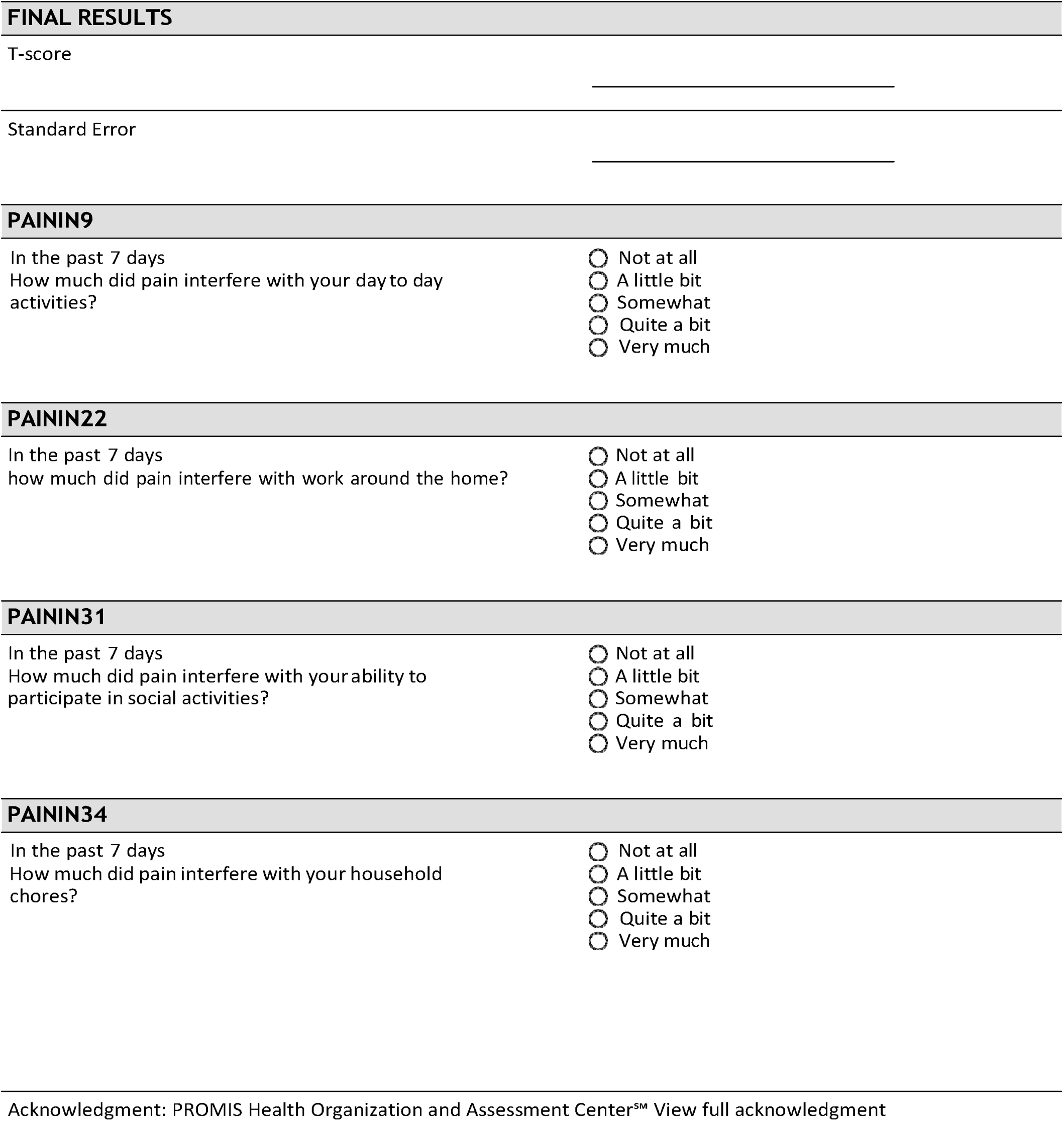

### Supplemental Text 3. Clinical Trial Evaluation Survey

#### Evaluation Survey

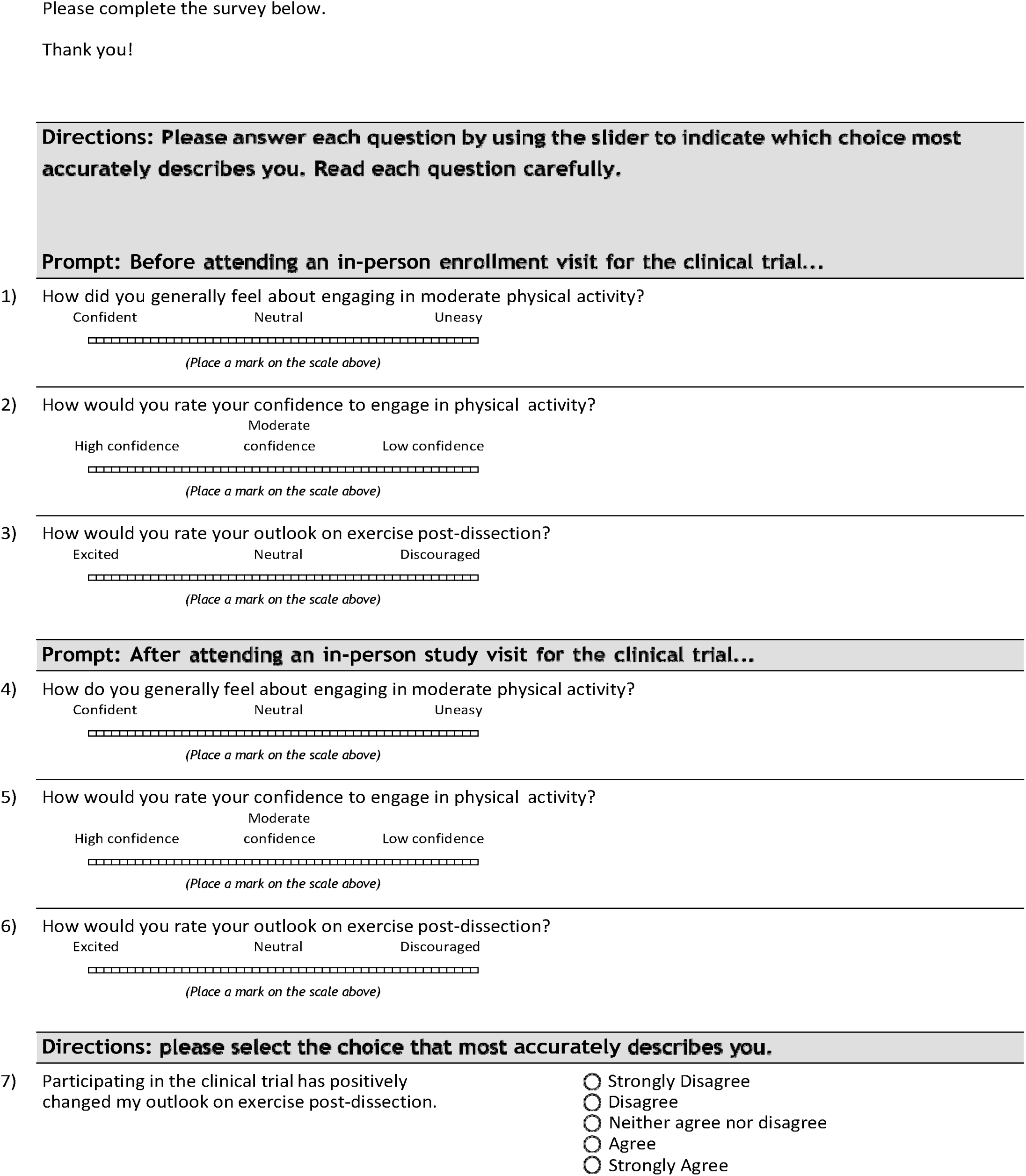
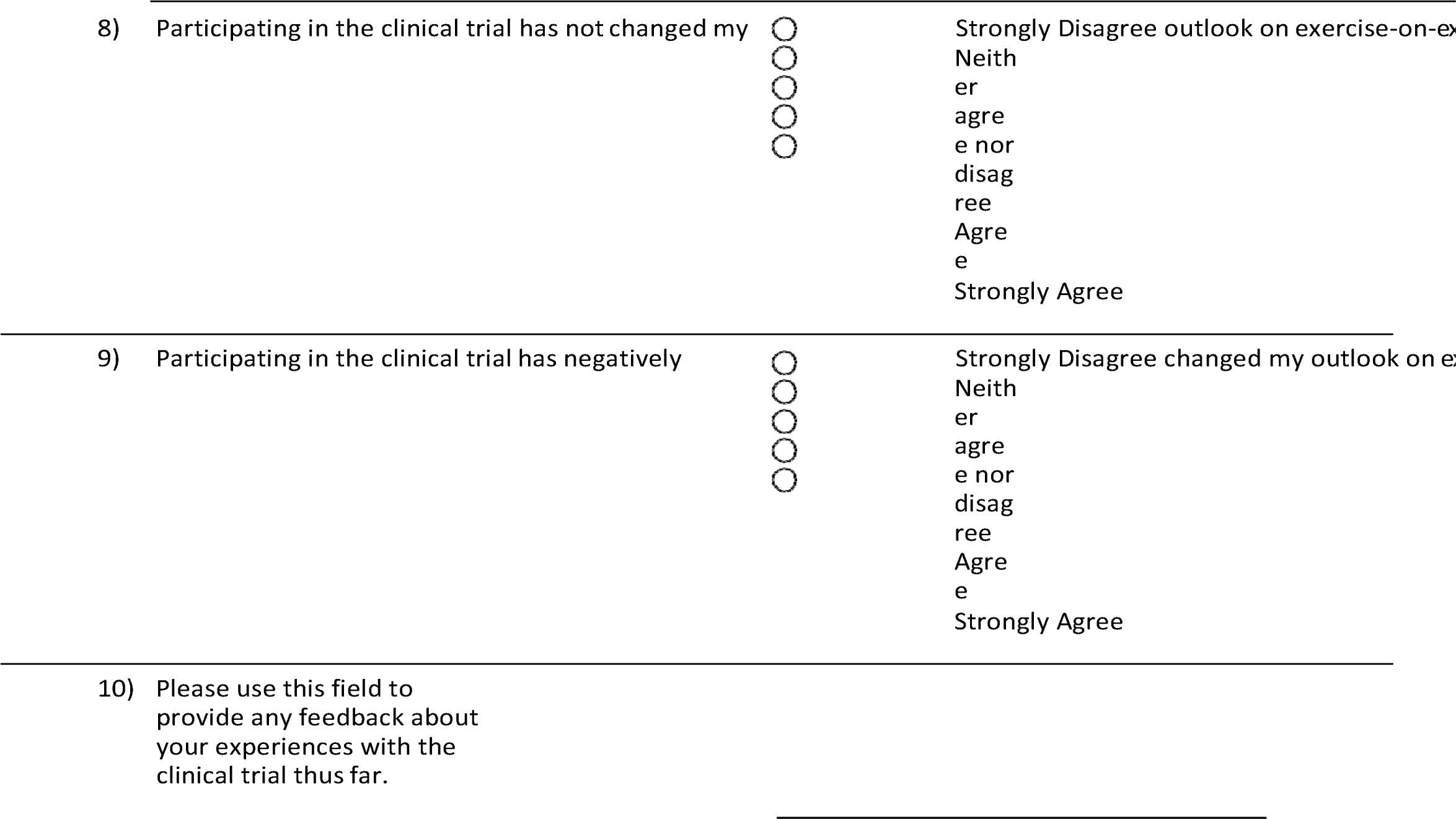

## Notes

### Competing Interest Statement

The authors have declared no competing interest.

### Clinical Trial

NCT05610462

### Author Declarations

The study protocol was reviewed and approved by the Committee for the Protection of Human Subjects at the University of Texas Health Science Center at Houston (UTHealth Houston), and institutional review boards at the University of Michigan and Washington University School of Medicine in St. Louis. All subjects signed a written informed consent document prior to enrollment.

